# Insulin-like growth factor 2 hypermethylation in peripheral blood leukocytes and colorectal cancer risk and prognosis: a propensity score analysis of multiple-centre populations

**DOI:** 10.1101/2022.04.27.22274374

**Authors:** HongRu Sun, YanLong Liu, YuXue Zhang, Yibaina Wang, BinBin Cui, YaShuang Zhao, YuPeng Liu

**Affiliations:** Department of Epidemiology, Public Health College, Harbin Medical University, 157 Baojian Street, Harbin 150081, Heilongjiang Province, China; HongRu Sun, Yibaina Wang, YaShuang Zhao, and YuPeng Liu; Department of Colorectal Surgery, The Third Affiliated Cancer Hospital of Harbin Medical University, 150 Haping Street, Harbin 150081, Heilongjiang Province, China; YanLong Liu and BinBin Cui; Department of Hygiene Microbiology, Public Health College, Harbin Medical University, 157 Baojian Street, Harbin 150081, Heilongjiang Province, China; YuXue Zhang

**Keywords:** IGF2 hypermethylation, colorectal neoplasms, risk, prognosis, propensity score analysis

## Abstract

**Background:** To comprehensively assess and validate the associations between insulin-like growth factor 2 (*IGF2*) gene methylation in peripheral blood leukocytes (PBLs) and colorectal cancer (CRC) risk and prognosis.

**Methods:** The association between *IGF2* methylation in PBLs and CRC risk was initially evaluated in a case-control study and then validated in a nested case-control study and a twins’ case-control study, respectively. Meanwhile, an initial CRC patient cohort was used to assess the effect of *IGF2* methylation on CRC prognosis and then the finding was validated in the EPIC-Italy CRC cohort and TCGA datasets. A propensity score (PS) analysis was performed to control for confounders, and extensive sensitivity analyses were performed to assess the robustness of our findings.

**Results:** PBL *IGF2* hypermethylation was associated with an increased risk of CRC in the initial study (OR_PS-adjusted_, 2.57, 95% CI: 1.65 to 4.03, *P*<0.0001), and this association was validated using two independent external datasets (OR_PS-adjusted_, 2.21, 95% CI: 1.28 to 3.81, *P*=0.0042 and OR_PS-adjusted_, 10.65, 95% CI: 1.26 to 89.71, *P*=0.0295, respectively). CRC patients with IGF2 hypermethylation in PBLs had significantly improved overall survival compared to those patients with IGF2 hypomethylation (HR_PS-adjusted_, 0.47, 95% CI: 0.29 to 0.76, *P*=0.0019). The prognostic signature was also observed in the EPIC-Italy CRC cohort, although the HR did not reach statistical significance (HR_PS-adjusted_, 0.69, 95% CI: 0.37 to 1.27, *P*=0.2359).

**Conclusions:** IGF2 hypermethylation may serve as a potential blood-based predictive biomarker for the identification of individuals at high risk of developing CRC and for CRC prognosis.

**Funding:** This work was supported by the China Postdoctoral Science Foundation (grant number 2018M641875 to YPL); the Natural Science Foundation of Heilongjiang Province (grant number YQ2019H021 to YPL); the National Natural Science Foundation of China (grant number 81473055 to YSZ), and by grant from the SCORE Foundation (Y-MX2016-045 to YLL).

**Clinical trial number:** Not applicable.

## 1. INTRODUCTION

Colorectal cancer (CRC) is the third-most common cancer in men and the second-most in women worldwide, with an estimated 1,931,590 newly diagnosed cases and 935,173 deaths in 2020, accounting for approximately 1 in 10 cancer cases and deaths.[1 2] In China, the number of CRC cases has rapidly increased since the 1980s, with an estimated 555,477 newly diagnosed patients and 286,162 deaths in 2020, accounting for approximately 30% of all annually diagnosed CRC cases and CRC-related deaths worldwide.[2 3] The initiation and progression of CRC is multifactorial and gradual with progressive accumulation of both genetic and epigenetic abnormalities, including aberrant DNA methylation and loss of imprinting (LOI).[4–6] The insulin-like growth factor 2 (*IGF2*) gene is one of the first imprinted genes identified in humans. The IGF2 protein has a tumour-promoting effect on existing colorectal neoplasia[7–10] and that LOI of IGF2 in either tissue or peripheral blood leukocyte (PBL) samples is associated with an increased risk of CRC.[11 12] Furthermore, a recent cohort study showed a significant association between *IGF2* hypomethylation in paraffin-embedded tissues and poor CRC prognosis.[13] However, no study has evaluated whether PBL IGF2 methylation, which can be determined using non-invasive techniques, is associated with CRC risk or prognosis.

We therefore performed this study to comprehensively assess the association between PBL IGF2 methylation status and CRC risk and prognosis. The propensity score (PS) method has been increasingly used to reduce the likelihood of confounding bias in observational studies. It is a powerful statistical tool to control for confounding bias and is often more practical and statistically more efficient than conventional strategies including covariate matching, stratified analysis, and multivariate statistical analysis.[14 15] In this study, we used PS-based methods to assess the effect of PBL IGF2 methylation on the risk of developing CRC, and then further validated our findings using external datasets from EPIC-Italy CRC cohort and GEO.[16–18] In addition, we used the same PS-based methods to assess the association between IGF2 methylation and CRC prognosis using CRC patient PBLs and tumour tissues, and then further validated our findings using external datasets from EPIC-Italy CRC cohort and The Cancer Genome Atlas (TCGA).[17 19]

## 2. METHODS

### 2.1. Study design and participants

#### 2.1.1. Initial study

##### 2.1.1.1. IGF2 methylation in PBLs and CRC risk

Descriptions of the study design and patient selection strategy have been published elsewhere.[20 21] Briefly, this hospital-based case-control study included 281 primary sporadic CRC patients diagnosed (from June 2004 to May 2005 and May 2007 to January 2008) at the Cancer Hospital of Harbin Medical University (HMU) and 147 CRC patients at the Second Affiliated Hospital of HMU (from October 2010 to December 2011) in Harbin, China (**Figure 1**). During the same time period, 428 cancer-free controls were selected from the Second Affiliated Hospital of HMU by individual matching on gender and age (±2 years). The basic characteristics of the CRC cases and controls are shown in **Supplementary File 1A**. All participants provided written informed consent prior to participation in the study. Blood samples were collected before surgery for the CRC patients and before any therapy for the controls. This study was approved by the Ethics Committee of HMU.

**Figure 1.**
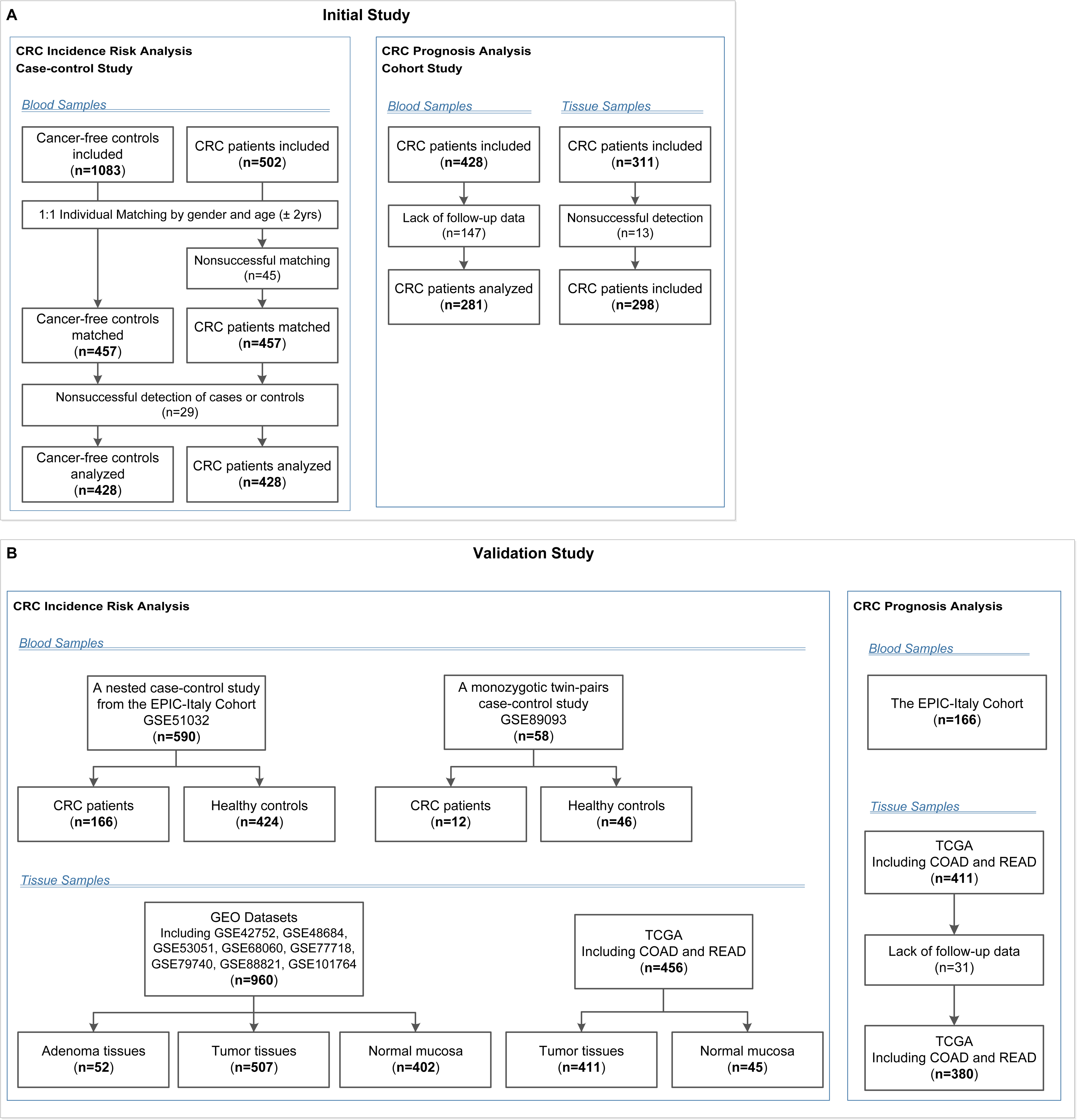
Flow chart of participants included and analysed in the (**A**) initial and (**B**) validation studies. COAD, colon adenocarcinoma; CRC, colorectal cancer; EPIC-Italy, the European Prospective Investigation into Cancer and nutrition (EPIC cohort) in Italy; GEO, Gene Expression Omnibus; READ, rectum adenocarcinoma; TCGA, The Cancer Genome Atlas.

All CRC patients had operable stage I-IV CRC, and their CRC diagnosis was histologically confirmed by a senior surgeon (YYL). Patients with adenomatous polyposis coli, who had a family history of CRC in first-degree relatives according to the Amsterdam criteria,[22] or who received anti-cancer therapy before surgery were excluded.

##### 2.1.1.2. IGF2 methylation in PBLs and CRC prognosis

The 281 CRC patients diagnosed at the Cancer Hospital of HMU were all included in the final CRC cohort, while the 147 CRC patients from the Second Affiliated Hospital of HMU were excluded because of the lack of follow-up information. For each patient, extensive demographic, clinicopathological and treatment information were extracted from the electronic medical record system. All surgical operations were performed by the same surgical oncologists (BBC and YLL) and all patients had negative surgical margins. The basic characteristics of the CRC patients included in this cohort are shown in **Supplementary File 1B**.

The primary outcomes were overall survival (OS) from CRC diagnosis to death and disease free survival (DFS) from CRC diagnosis to disease recurrence, metastasis, or death from CRC, whichever came first. Another outcome was CRC-specific survival (CSS), which was defined as the time from disease diagnosis to CRC-specific death. Outcomes were observed during the follow-up period through March 15, 2014 via an established protocol. Postoperative patients were followed up at 3-6 months intervals for the first year and then annually. We used a telephone-delivery follow-up questionnaire to collect information on the date and cause of death of the CRC patients. The recorded date and cause of death of the CRC patients were validated using the medical certification of death and the Harbin death registration system. Of the 281 eligible CRC patients included in this analysis, 127 died, 120 remained alive and 34 were lost to follow-up.

##### 2.1.1.3. IGF2 methylation in tumour tissues and CRC prognosis

Fresh tumour tissues were collected during surgery from the CRC patients treated at the Cancer Hospital of HMU, with written consent being obtained prior to surgery. Among the 298 eligible CRC patients included in this analysis, 124 died, 141 remained alive and 32 were lost to follow-up. In all, 185 paired tumour tissue and PBL samples were obtained from the same patients.

##### 2.1.1.4. Sample size

The sample size was estimated using PASS version 11.0.7 (NCSS LLC., USA). To assess whether aberrant methylation of insulin-like growth factor 2 (IGF2) in peripheral blood leukocytes (PBLs) is associated with the risk of developing colorectal cancer (CRC) in our present case-control study, we estimated the sample size according to a logistic regression model. A sample size of 652 participants was needed to achieve 90% power (at the 5% level of statistical significance) in order to detect an odds ratio (OR) of 1.8 or more with a 20% prevalence in the control group. In addition, taking into consideration incomplete questionnaires and the failure rate for MS-HRM detection, we included approximately 20% more patients and finally targeted a total sample size of 800 participants (n = 400 for CRC patients and controls, respectively).

To assess whether aberrant IGF2 methylation in PBLs is associated with CRC patient prognosis in our present cohort study, we estimated the sample size according to a Cox regression model. A sample size of 184 CRC patients was needed to achieve 80% power (at the 5% level of statistical significance) to detect a hazard ratio (HR) of 0.5 or 2.0 with an overall event rate of 50% in this cohort. In addition, taking into consideration incomplete questionnaires and the failure rate for MS-HRM detection, we included approximately 20% more patients and finally targeted a total sample size of 221 CRC patients.

#### 2.1.2. Validation study

A case-control study nested in the EPIC-Italy cohort (GSE51032) and a twins’ case-control study (GSE89093) from the GEO were used to validate the association between PBL IGF2 methylation and CRC risk. We also used the EPIC-Italy CRC cohort and TCGA datasets (COAD and READ) to validate the relationship between IGF2 methylation in PBLs or tumour tissues and CRC prognosis, respectively. The basic characteristics of validation populations are shown in **Supplementary File 1**.

### 2.2. Methylation analysis

Sample processing, DNA extraction and bisulfite modification were performed as described previously.[20] Briefly, DNA was extracted from buffy coats and tumour tissues using a QIAamp DNA Blood Mini Kit (Qiagen, Hilden, Germany, Cat#51106) and a DNeasy Blood & Tissue Kit (Qiagen, Cat #69506), respectively. The DNA was then bisulfite-modified using an EpiTect Plus DNA Bisulfite Kit (Qiagen, Cat#59826) according to the manufacturer’s protocols. The bisulfite-modified DNA sample was quantified using a NanoDrop 2000c bioanalyzer (Thermo-Fisher, USA), diluted to a final concentration of 10 ng/µL and divided into aliquots for storage (−20 °C).

We designed a methylation-sensitive high-resolution melting (MS-HRM) assay for the IGF2 promotor region (101bp, GRCh38/hg38; chr11:2139870-2139971, including 11 CpG sites, **Figure 2**) using Methprimer,[23] and IGF2 methylation status was tested with the researchers being blinded to patient outcome. A set of methylation standards (100, 25, 10, 5, 2, 1, and 0% methylated DNA) were prepared by mixing commercially available methylated and unmethylated DNA (Zymo Research, Irvine, USA, Cat#D5014); these standards were used to semi-quantitatively measure the methylation level of the IGF2 target region in the samples. The MS-HRM analysis was performed as previously described. Briefly, each PCR mixture had a total volume of 10 µL and contained 2× LightCycler 480 High Resolution Melting Master Mix (Roche Applied Science, Mannheim, Germany, Cat#4909631001), 3 mmol/L MgCl2, 0.4 µmol/L of each primer (forward primer: GGGATTTGGTTGAGGTTTTAAG; reverse primer: TACGACTAAAAAAACCCCTAAACTC) and 1 µL (approximately 10 ng) of bisulfite-modified template DNA. PCR conditions were as follows: initial PCR activation (95 °C for 15 minutes); 55 3-step cycles (95 °C for 10 seconds, 56 °C for 30 seconds, and 72 °C for 20 seconds); and final extension (72 °C for 10 minutes). A blank control (non-template control) sample was included in each batch, and all reactions were performed in duplicate (as technical replicates). A third trial was conducted for the samples that presented inconsistent results between the two trials. PCR amplification and MS-HRM analyses were performed using the LightCycler 480 platform (Roche). After normalization of the melting curves using the software module of Gene Scanning (Roche), two investigators (YP.L. and HR.S.), who were blinded to the sample groups, independently assessed the MS-HRM data. Discrepancies were resolved by discussion and consensus with another investigator (YBN.W.).

**Figure 2.**
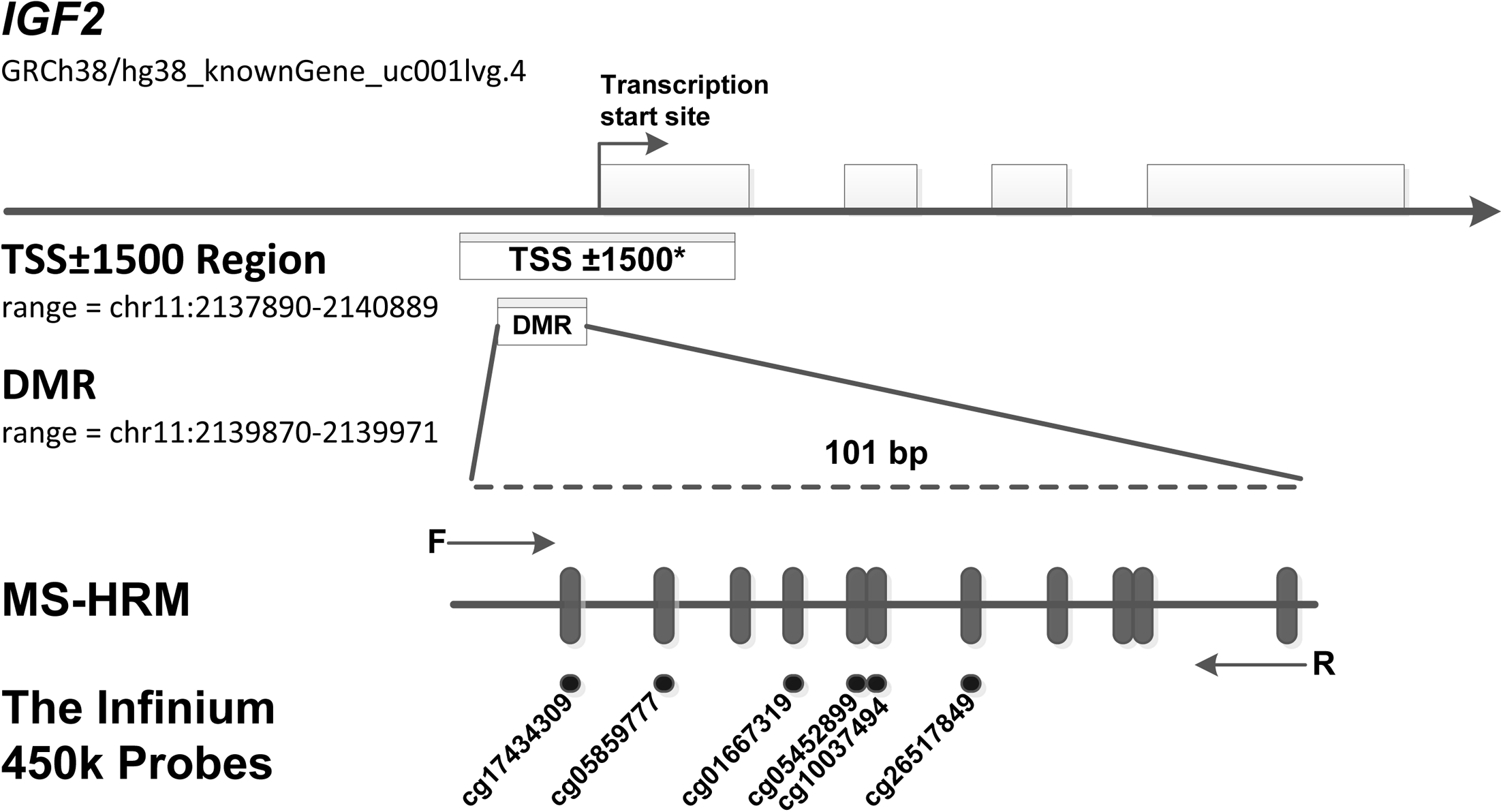
MS-HRM assay to detect methylation levels at the IGF2 promoter region. Our assay encompassed a 101 bp region (range: chr11: 2139870-2139971, including 11 CpGs), which located in the promoter of human IGF2 gene and near the transcription start site (TSS). Arrows indicate position and direction of MS-HRM primers. **Black dashes** (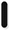) indicate individual CpGs on DMR region tested in our study. **Black Dots** (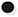) indicate six methylation probes (including cg17434309, cg05859777, cg01667319, cg05452899, cg10037494, cg26517849) on the Illumina Infinium HumanMethylation450 BeadChip, which overlap CpG sites detected in our DMR region. *The TSS±1500 region is an expanded region of −1500 bp upstream to +1500 downstream from the TSS (range: chr11: 2137890-2140889). To test the robustness of our results, we further assess the association of methylation levels of this expanded region with CRC risk. In the TSS±1500 region, a total of 32 CpG sites were detected by the Infinium HumanMethylation450 methylation probes. The methylation probes are as follows: cg25163476, cg24917382, cg24366657, cg23905216, cg23676551, cg22287492, cg21667878, cg20339650, cg19443075, cg19371526, cg19002337, cg18087943, cg17434309, cg17037101, cg16415340, cg15393937, cg14608156, cg14188639, cg13756879, cg12773325, cg12614029, cg11915650, cg10659464, cg10037494, cg09694722, cg08162473, cg05859777, cg05452899, cg04072545, cg03760951, cg02835822, cg01667319.

### 2.3. Covariates, Missing data analysis and imputation

All participants were interviewed face-to-face to complete a structured standard questionnaire, which was partially adopted from Shu et al.[24] The questionnaire queried information on demographic characteristics, lifestyle factors (including family history, smoking, alcohol consumption, occupational physical activity), and diet. Dietary consumption over the past year was assessed using a validated food frequency questionnaire (FFQ).[24] The FFQ included nine major food groups, which represented most of the common foods in Northeast China. The food items included barbecued foods, coarse grains, fish stewed with brown sauce, fresh fruits, fried foods, green vegetables, leftovers and pork.

Questionnaire-derived covariates included: age (< 60, ≥ 60), gender (male, female), BMI (< 24, ≥ 24), family history of cancer other than CRC in first-degree relatives (no, yes), occupational physical activity (blue-collar, white-collar), smoking status (no, yes), and consumption of barbecued foods (< 1, ≥ 1 times/week), coarse grains (< 50, ≥ 50 g/week), fish stewed with brown sauce (< 1, ≥ 1 times/week), fruits (< 2, ≥ 2 times/week), fried foods (< 1, ≥ 1 times/month), green vegetables (< 100, ≥ 100 g/day), leftovers (< 1, ≥ 1 times/week) and pork (< 250, ≥ 250 g/week). All questionnaire-derived variables were analysed via missing value analysis and were imputed via multiple imputations as described in our previous study.[20]

### 2.4. Statistical analysis

Means and standard deviations were reported for continuous variables, while counts and frequencies were reported for categorical variables. Covariate differences between groups were compared using the standardised differences method with a significant imbalance level of standardised difference ≥ 25%. In the CRC risk analysis, we first categorised individuals into two groups according to the optimal cut-off point for the IGF2 methylation level (≤ 1% hypomethylation group; > 1% hypermethylation group), which was determined using the receiver operating characteristic (ROC) curve and the Youden index method with case-control status as the dependent variable (0 for controls; 1 for cases). We then conducted univariate and multivariate logistic regression analyses and reported odd ratios (ORs) and 95% confidence intervals (CIs) to assess the association between IGF2 methylation status and CRC risk. In the CRC prognosis analysis, the cut-off point for the IGF2 methylation level was also 1% using the same method with overall survival time as the dependent variable (0 for less than median survival time; 1 for longer than or equal to median survival time). According to this cut-off point, CRC patients were categorised into IGF2 hypomethylation and IGF2 hypermethylation groups (206 and 75 cases, respectively). Kaplan-Meier curves and log-rank tests were used to compare OS, DFS or CSS between groups. The associations between IGF2 methylation and OS, DFS or CSS were estimated using univariate and multivariate Cox regression models and reported as hazard ratios (HRs) and 95% CIs. Two-sided statistical significance was defined as *P*<0.05. ROC analyses were performed with MedCalc version 15.4 (Ostend, Belgium) and all other statistical analyses were performed with SPSS Statistics version 23.0 (IBM, Inc., USA).

To minimise group differences on covariates, we performed a PS-based analysis. In the CRC risk analysis, the PS was calculated with case-control status as the dependent variable using a multivariate logistic regression model that included demographic and lifestyle factors (**Supplementary File 2A**). In the survival analysis, the PS was calculated with IGF2 methylation as the dependent variable using a multivariate logistic regression model that included demographic and lifestyle factors and clinicpathological characteristics (**Supplementary File 2B**). To incorporate all the patients in the analyses, we primarily employed the PS-adjustment method.[14] Additionally, several PS-based methods, including stratification by quintile of PS, weighting with inverse probability of treatment weights, and individual PS matching, were also performed as sensitivity analyses.

For PS-stratification analyses, all patients were stratified by PS quintile. HRs for PS-stratified analyses were obtained by pooling effect estimates from each PS quintile. For PS-weighted analyses, each patient was weighted by the inverse probability of being in the higher versus lower IGF2 methylation groups; this method is known as weighting with inverse probability of treatment weights (IPTW). The weight for each patient was calculated according to the method described by Robins and Hernan:[25] Pt/PS and (1-Pt)/(1-PS) for patients in the IGF2 hypermethylation and hypomethylation groups, respectively; where Pt is the proportion of the patients with IGF2 hypermethylation in all participants. In addition to using PS as an adjustment or weighting factor in the analysis model, PS was also used for individual matching. PS matching was performed using a nearest-neighbour algorithm with a calliper of 0.2 (which means that the maximum allowable PS difference between groups was no larger than 20% of the standard deviation of PS).

### 2.5. Sensitivity analysis

To assess the robustness of our results, we performed extensively predefined sensitivity analyses. First, to evaluate the potential impact of the PS-adjusted confounders on our results in the initial study, we performed confounding RR analysis, which was defined as the ratio of the PS-adjusted effect estimates and the unadjusted effect estimates.[26] To investigate whether potential residual confounders could impact the results, we calculated the E-value for PS-adjusted effect estimates and the limit of the CI closest to the null.[27] Finally, we performed subgroup analyses according to the tumour location (colon or rectum), UICC stage, gender (female vs. male), age (≥60 vs. <60 years), and body mass index (BMI, ≥24 vs. <24).

Additionally, we performed several post hoc sensitivity analyses. To test whether tumour load impacts IGF2 methylation level in PBLs, we assessed the associations between IGF2 methylation and UICC stage and serum carcinoembryonic antigen (CEA) level. We also tested whether PBL IGF2 methylation levels could have been impacted by leukocyte count and the percentage of certain subpopulations. In addition, to comprehensively determine the association between IGF2 methylation and CRC risk, we combined the results from the initial case-control and the validation studies using a meta-analysis method of random effect model. Finally, we explored whether PBL IGF2 methylation levels differ between the CRC patients included in the survival analysis and those excluded.

## 3. RESULTS

### 3.1. IGF2 methylation in PBLs and CRC risk

As shown in **Figure 3A**, subjects with IGF2 hypermethylation (15.54%), compared to subjects with IGF2 hypomethylation (84.46%), had a significantly increased CRC risk (PS-adjusted OR, 2.57, 95% CI: 1.64-4.03; *P*<0.001). This association remained statistically significant even after subgroup analyses. Notably, stratified analyses showed a positive association only in UICC stage I-III but not in stage IV cancers.

**Figure 3.**
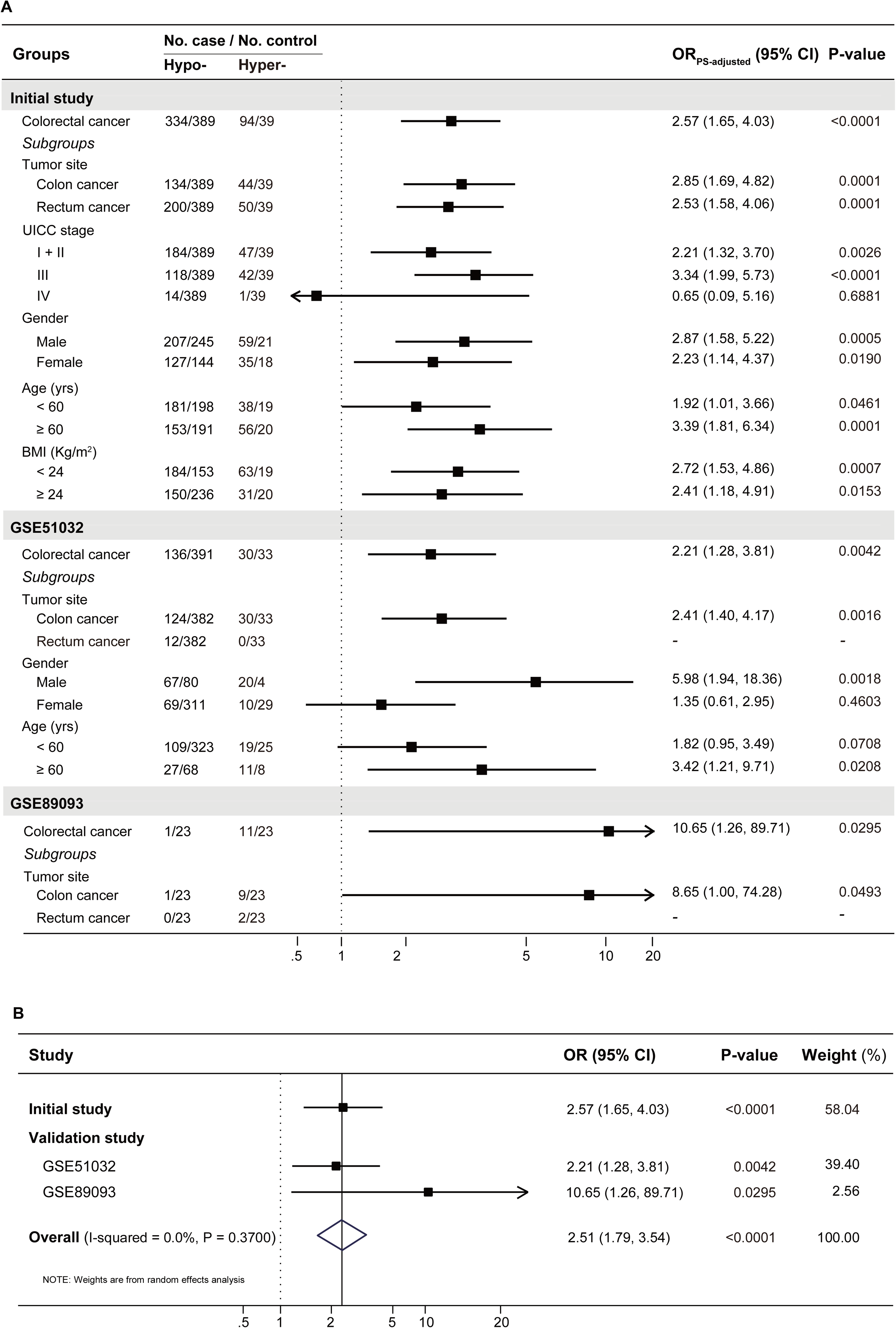
The results for the association between PBL IGF2 methylation and the risk of developing CRC (**A**) in the initial and validation studies, and (**B**) in the combined populations. *A total of 14 covariates were included in the propensity score model in the initial case-control study (see supplementary table S3). In the validation studies, gender and age were included in the the propensity score model for GSE51032 and age for GSE89093, respectively. To be conservative, we pooled the results from these three datasets with a random effect model in the meta-analysis. BMI, body mass index; CI, confidence interval; Hyper-, IGF2 hypermethylation; Hypo-, IGF2 hypomethylation; OR, odds ratio.

In the EPIC-Italy dataset, IGF2 hypermethylation was also significantly associated with an increased CRC risk (OR, 2.21, 95% CI: 1.28-3.81; *P*=0.004). Subgroup analyses showed the association was statistically significant in distal colon cancer, men, and older subjects (**Figure 3A**). Using the GSE89093 dataset, IGF2 hypermethylation was strongly associated with an increased risk of CRC or colon cancer alone. After pooling the results from these three datasets, the observed association was still significant (**Figure 3B**).

### 3.2. IGF2 methylation in PBLs and CRC prognosis

The Kaplan-Meier survival curves for the PBL IGF2 hypermethylation and hypomethylation groups are shown in **Figure 4**. The median OS was 73 months (95% CI: 66-80 months) in the IGF2 hypermethylation group versus 56 months (95% CI: 52-61 months) in the hypomethylation group. The OS rate was 75% (95% CI: 60-80%) in the IGF2 hypermethylation group versus 62% (95% CI: 50-57%) in the hypomethylation group (HR_PS-adjusted_, 0.47, 95% CI: 0.29-0.76; *P*=0.002). The CSS rate was 56% (95% CI: 45-68%) in the IGF2 hypermethylation group versus 38% (95% CI: 31-45%) in the hypomethylation group (HR_PS-adjusted_, 0.49, 95% CI: 0.30-0.80; *P*=0.004). The median DFS was 66 months (95% CI: 57-74 months) in the IGF2 hypermethylation group versus 52 months (95% CI: 48-57 months) in the hypomethylation group (HR_PS-adjusted_, 0.53, 95% CI: 0.32-0.85; *P*=0.009). Based on subgroup analyses, we found that the statistically significant association persisted in rectal cancers, UICC stage I-III cancers, males, older, or normal body weight patients. In contrast, the effect estimates did not reach statistical significance in colon cancers, stage IV cancers, females, younger, and overweight or obese patients (**Table 1**).

**Figure 4.**
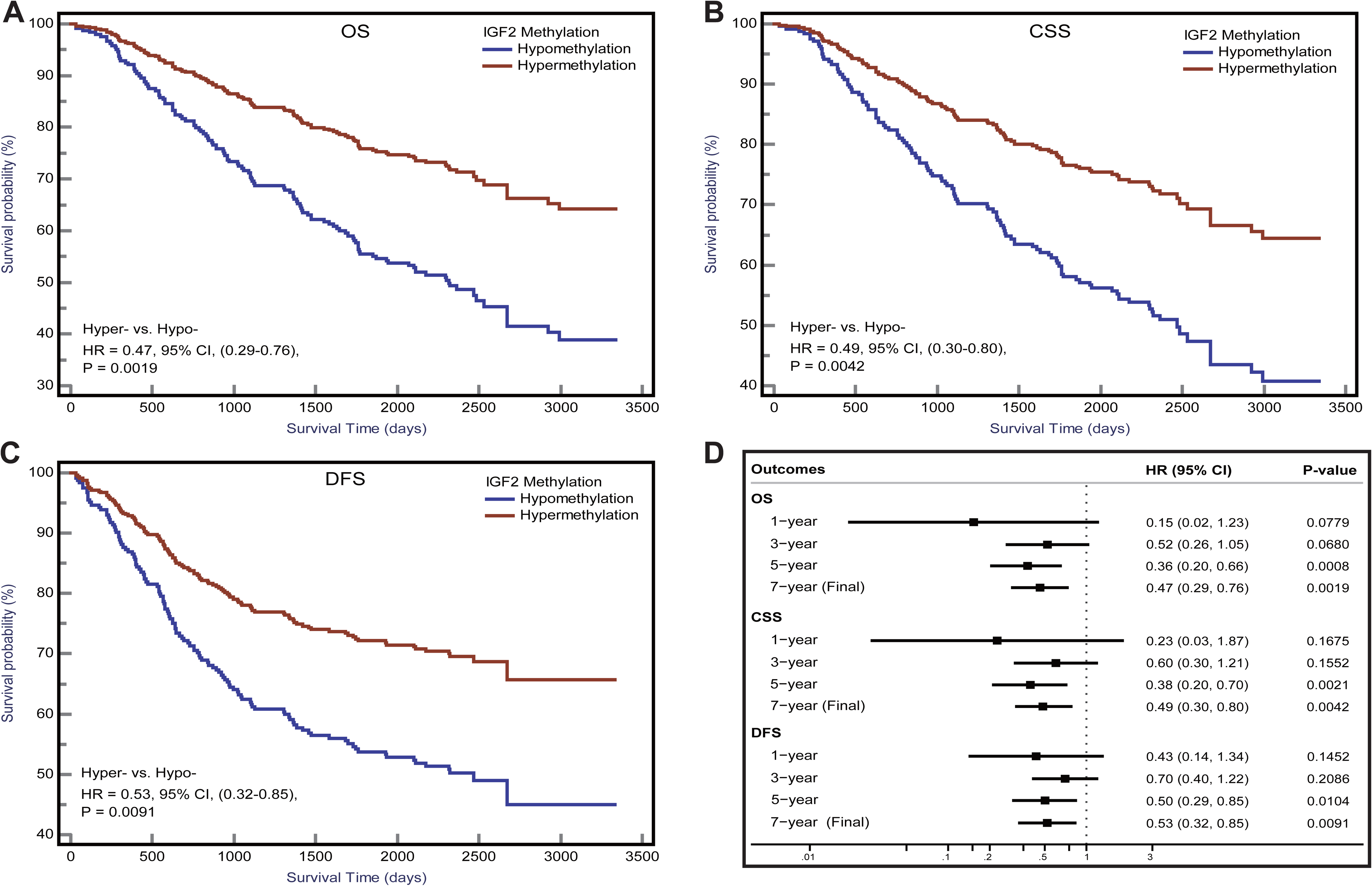
IGF2 methylation in PBLs and CRC prognosis in the initial cohort. Kaplan-Meier plots for (**A**) overall survival, (**B**) CRC specific survival, and (**C**) disease-free survival according to IGF2 methylation status in PBLs in CRC patients. (**D**) Associations of *IGF2* methylation in PBLs with CRC prognosis during different follow-up periods. CI, confidence interval; CRC, colorectal cancer; CSS, cancer specific survival; DFS, disease free survival; HR, hazard ratio; Hyper-, IGF2 hypermethylation; Hypo-IGF2 hypomethylation; OS, overall survival; PBL, peripheral blood leukocyte.

**Table 1.**
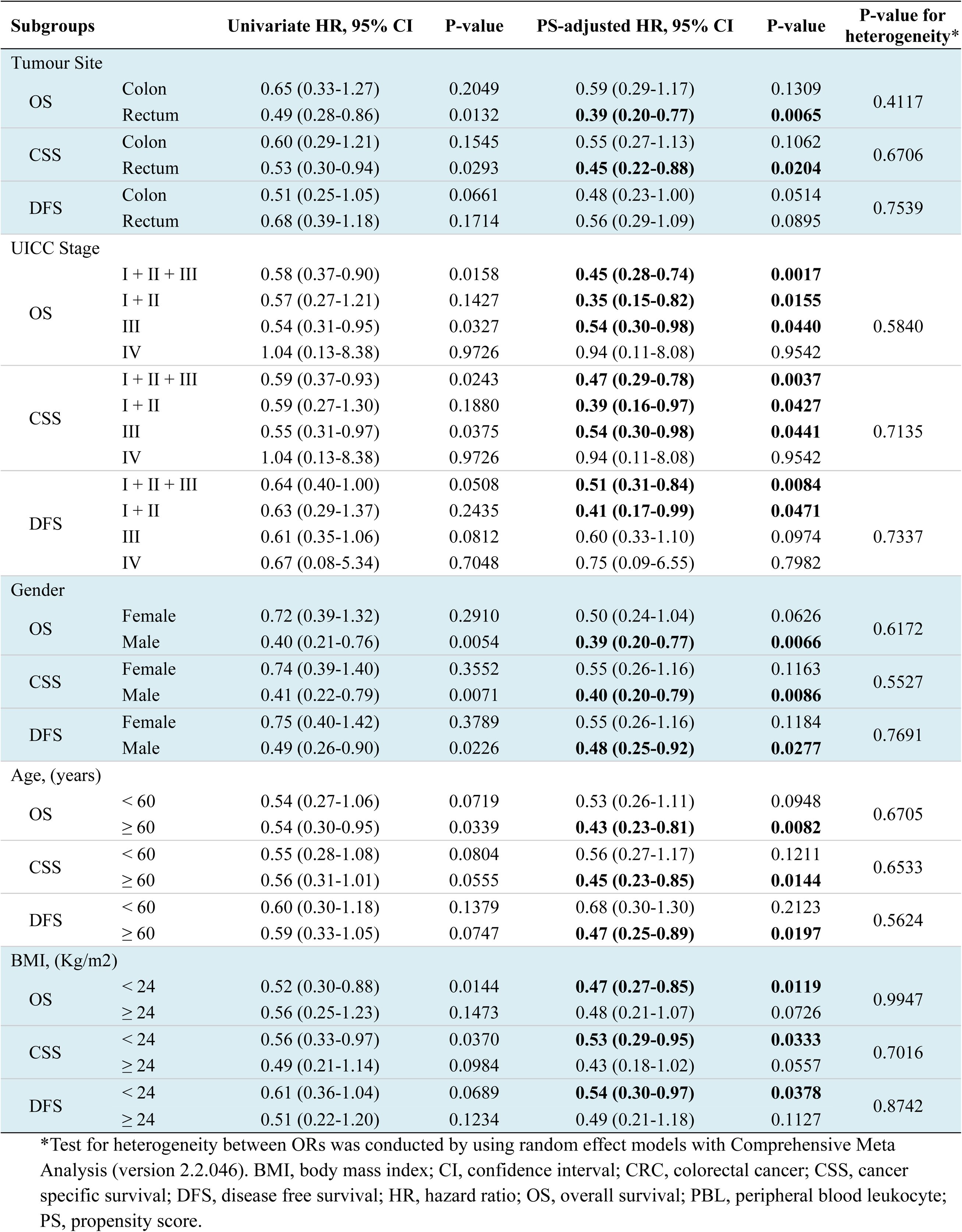
Subgroup analysis for the association of PBL IGF2 methylation with CRC prognosis.

| Subgroups |  | Univariate HR, 95% CI | P-value | PS-adjusted HR, 95% CI | P-value | P-value for heterogeneity* |
| --- | --- | --- | --- | --- | --- | --- |
| Tumour Site |  |  |  |  |  |  |
| OS | Colon | 0.65 (0.33-1.27) | 0.2049 | 0.59 (0.29-1.17) | 0.1309 | 0.4117 |
|  | Rectum | 0.49 (0.28-0.86) | 0.0132 | **0.39 (0.20-0.77)** | **0.0065** |  |
| CSS | Colon | 0.60 (0.29-1.21) | 0.1545 | 0.55 (0.27-1.13) | 0.1062 | 0.6706 |
|  | Rectum | 0.53 (0.30-0.94) | 0.0293 | **0.45 (0.22-0.88)** | **0.0204** |  |
| DFS | Colon | 0.51 (0.25-1.05) | 0.0661 | 0.48 (0.23-1.00) | 0.0514 | 0.7539 |
|  | Rectum | 0.68 (0.39-1.18) | 0.1714 | 0.56 (0.29-1.09) | 0.0895 |  |
| UICC Stage |  |  |  |  |  |  |
| OS | I + II + III | 0.58 (0.37-0.90) | 0.0158 | **0.45 (0.28-0.74)** | **0.0017** | 0.5840 |
|  | I + II | 0.57 (0.27-1.21) | 0.1427 | **0.35 (0.15-0.82)** | **0.0155** |  |
|  | III | 0.54 (0.31-0.95) | 0.0327 | **0.54 (0.30-0.98)** | **0.0440** |  |
|  | IV | 1.04 (0.13-8.38) | 0.9726 | 0.94 (0.11-8.08) | 0.9542 |  |
| CSS | I + II + III | 0.59 (0.37-0.93) | 0.0243 | **0.47 (0.29-0.78)** | **0.0037** | 0.7135 |
|  | I + II | 0.59 (0.27-1.30) | 0.1880 | **0.39 (0.16-0.97)** | **0.0427** |  |
|  | III | 0.55 (0.31-0.97) | 0.0375 | **0.54 (0.30-0.98)** | **0.0441** |  |
|  | IV | 1.04 (0.13-8.38) | 0.9726 | 0.94 (0.11-8.08) | 0.9542 |  |
| DFS | I + II + III | 0.64 (0.40-1.00) | 0.0508 | **0.51 (0.31-0.84)** | **0.0084** | 0.7337 |
|  | I + II | 0.63 (0.29-1.37) | 0.2435 | **0.41 (0.17-0.99)** | **0.0471** |  |
|  | III | 0.61 (0.35-1.06) | 0.0812 | 0.60 (0.33-1.10) | 0.0974 |  |
|  | IV | 0.67 (0.08-5.34) | 0.7048 | 0.75 (0.09-6.55) | 0.7982 |  |
| Gender |  |  |  |  |  |  |
| OS | Female | 0.72 (0.39-1.32) | 0.2910 | 0.50 (0.24-1.04) | 0.0626 | 0.6172 |
|  | Male | 0.40 (0.21-0.76) | 0.0054 | **0.39 (0.20-0.77)** | **0.0066** |  |
| CSS | Female | 0.74 (0.39-1.40) | 0.3552 | 0.55 (0.26-1.16) | 0.1163 | 0.5527 |
|  | Male | 0.41 (0.22-0.79) | 0.0071 | **0.40 (0.20-0.79)** | **0.0086** |  |
| DFS | Female | 0.75 (0.40-1.42) | 0.3789 | 0.55 (0.26-1.16) | 0.1184 | 0.7691 |
|  | Male | 0.49 (0.26-0.90) | 0.0226 | **0.48 (0.25-0.92)** | **0.0277** |  |
| Age, (years) |  |  |  |  |  |  |
| OS | < 60 | 0.54 (0.27-1.06) | 0.0719 | 0.53 (0.26-1.11) | 0.0948 | 0.6705 |
|  | ≥ 60 | 0.54 (0.30-0.95) | 0.0339 | **0.43 (0.23-0.81)** | **0.0082** |  |
| CSS | < 60 | 0.55 (0.28-1.08) | 0.0804 | 0.56 (0.27-1.17) | 0.1211 | 0.6533 |
|  | ≥ 60 | 0.56 (0.31-1.01) | 0.0555 | **0.45 (0.23-0.85)** | **0.0144** |  |
| DFS | < 60 | 0.60 (0.30-1.18) | 0.1379 | 0.68 (0.30-1.30) | 0.2123 | 0.5624 |
|  | ≥ 60 | 0.59 (0.33-1.05) | 0.0747 | **0.47 (0.25-0.89)** | **0.0197** |  |
| BMI, (Kg/m2) |  |  |  |  |  |  |
| OS | < 24 | 0.52 (0.30-0.88) | 0.0144 | **0.47 (0.27-0.85)** | **0.0119** | 0.9947 |
|  | ≥ 24 | 0.56 (0.25-1.23) | 0.1473 | 0.48 (0.21-1.07) | 0.0726 |  |
| CSS | < 24 | 0.56 (0.33-0.97) | 0.0370 | **0.53 (0.29-0.95)** | **0.0333** | 0.7016 |
|  | ≥ 24 | 0.49 (0.21-1.14) | 0.0984 | 0.43 (0.18-1.02) | 0.0557 |  |
| DFS | < 24 | 0.61 (0.36-1.04) | 0.0689 | **0.54 (0.30-0.97)** | **0.0378** | 0.8742 |
|  | ≥ 24 | 0.51 (0.22-1.20) | 0.1234 | 0.49 (0.21-1.18) | 0.1127 |  |

\*Test for heterogeneity between ORs was conducted by using random effect models with Comprehensive Meta Analysis (version 2.2.046). BMI, body mass index; CI, confidence interval; CRC, colorectal cancer; CSS, cancer specific survival; DFS, disease free survival; HR, hazard ratio; OS, overall survival; PBL, peripheral blood leukocyte; PS, propensity score.

Using the follow-up results of the 166 CRC patients from the EPIC-Italy cohort, we observed a clear trend for a longer OS related with IGF2 hypermethylation, although the association was not statistically significant. The univariate HR was 0.65 (95% CI: 0.35-1.19; *P*=0.164;), and after PS adjustment, the HR_PS-adjusted_ was 0.69 (95% CI: 0.37-1.27; *P*=0.236; K-M survival curves are shown in **Figure 5**).

**Figure 5.**
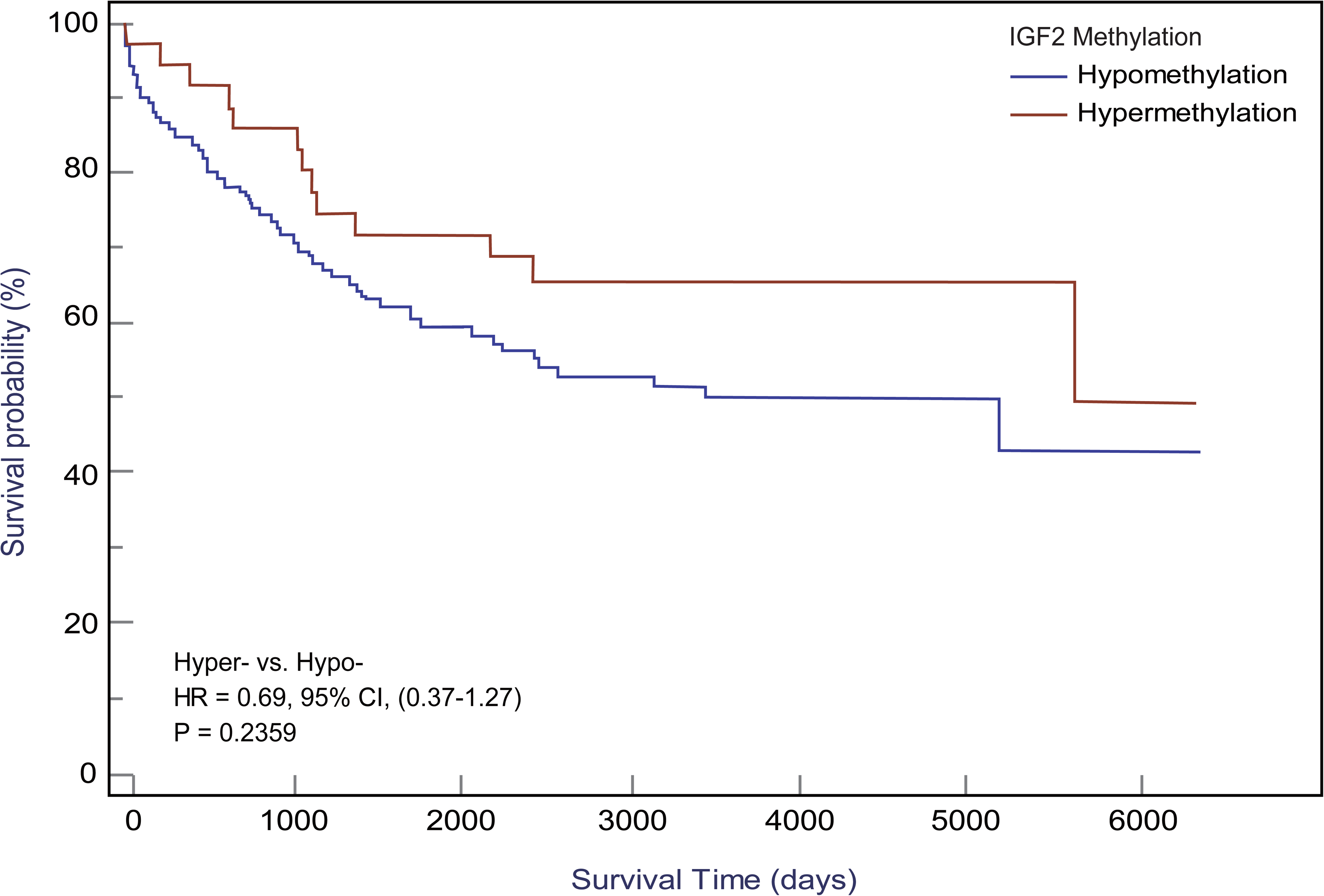
Kaplan-Meier curves for IGF2 methylation in PBLs and CRC overall survival in the EPIC-Italy cohort. CI, confidence interval; HR, hazard ratio; Hyper-, IGF2 hypermethylation; Hypo-IGF2 hypomethylation.

### 3.3. IGF2 methylation in tissues and CRC prognosis

In our initial study, the association between IGF2 methylation in tumour tissues and CRC patient survival did not reach statistical significance (**Figure 6A**), and this result was confirmed by the TCGA dataset (**Figure 6B**). Using IGF2 mRNA expression data in tumour tissues, we found an obvious negative correlation with IGF2 methylation (r=-0.24, *P*<0.001; **Figure 6C**). So we further explored whether IGF2 mRNA expression levels impact the disease prognosis but found no statistically significant association (**Figure 6D**).

**Figure 6.**
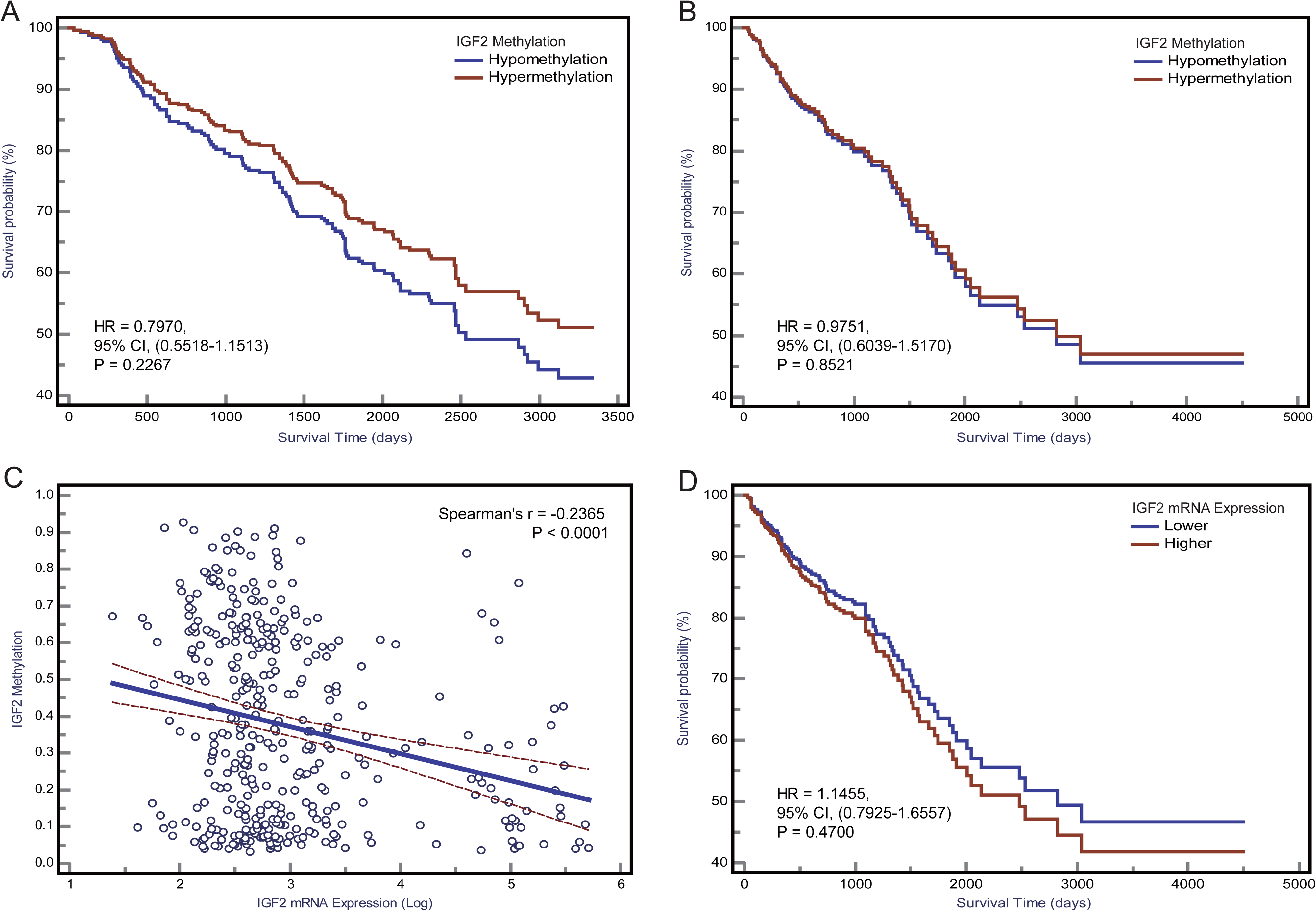
The association between IGF2 methylation in tumour tissues and CRC patient prognosis. Kaplan-Meier curves for overall survival according to IGF2 methylation status in tumour tissues of CRC patients in (**A**) the initial cohort and (**B**) TCGA datasets. (**C**) The correlation between IGF2 mRNA expression level and methylation level of the differentially methylated region of IGF2 tested in our study using the TCGA dataset. (**D**) Kaplan-Meier curves for overall survival according to IGF2 mRNA expression levels in tumour tissues of CRC patients in the TCGA cohort. CI, confidence interval; CRC, colorectal cancer; HR, hazard ratio.

### 3.4. Sensitivity analysis

We repeated analyses using other PS-based methods and the results were consistent with the PS-adjustment results (**Supplementary File 3**). Based on the confounding RR analysis (**Figure 7**), we did not find any substantial differences between the PS-adjusted effect estimates and the corresponding unadjusted effect estimates. Furthermore, the E-value analysis showed that our findings in both the initial and validation studies appear to be very robust (**Supplementary File 4**).

**Figure 7.**
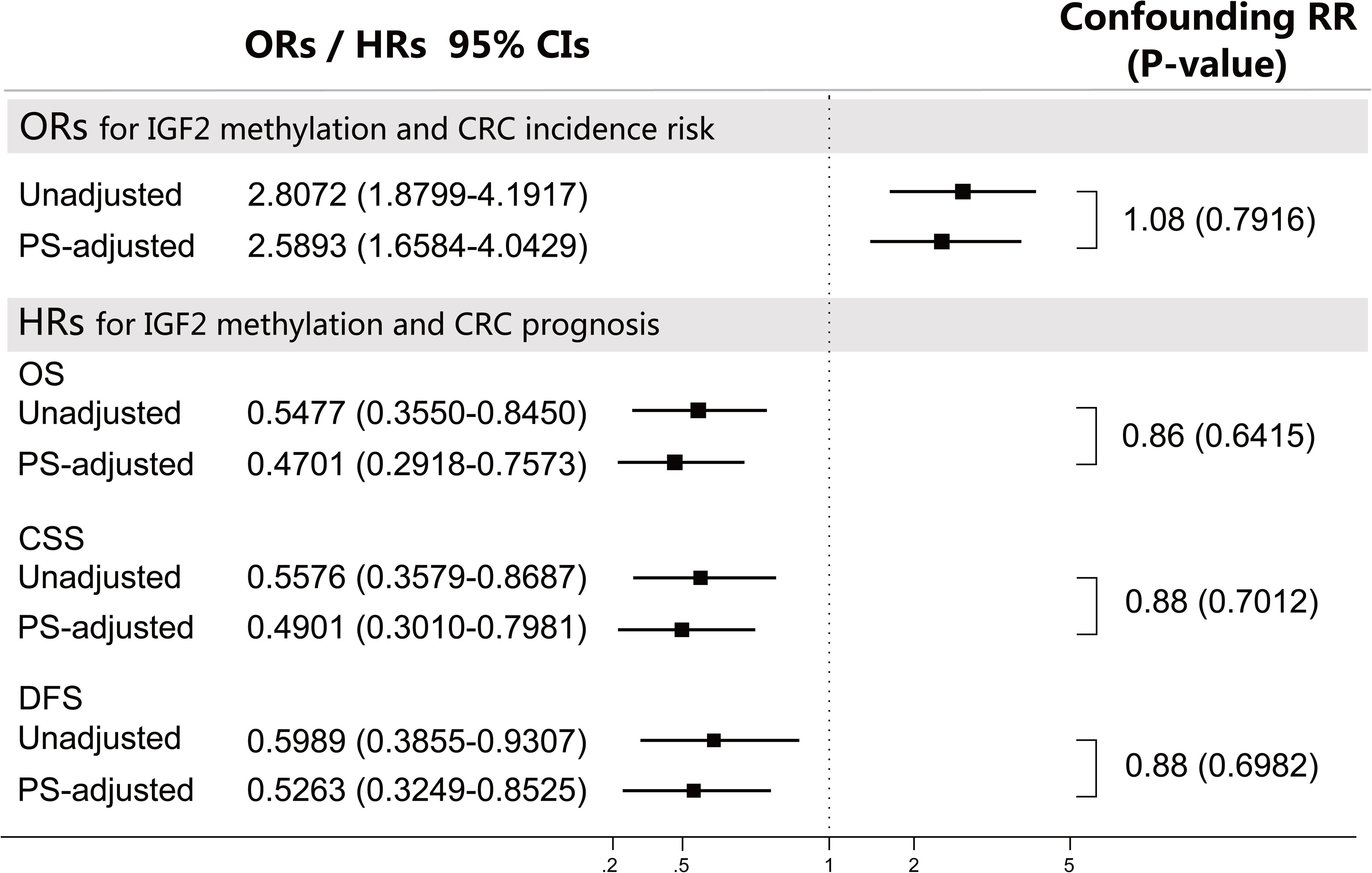
Sensitivity analyses using the confounding RR method. The confounding RR was defined as the ratio of the PS-adjusted effect estimates and the unadjusted effect estimates. CI, confidence interval; CRC, colorectal cancer; CSS, cancer specific survival; DFS, disease free survival; HR, hazard ratio; OR, odds ratio; OS, overall survival; PS, propensity score.

We found no evidence for an obvious impact of UICC stages on IGF2 methylation in PBLs. Of note, IGF2 hypermethylation was significantly more frequent in the CEA-low group than in the CEA-high group (34.15% vs. 20.89%, *P*=0.013). We found no significant relationship between the PBL IGF2 methylation levels and the leukocyte count or the percentages of certain leukocyte subpopulations. Finally, we analysed samples from 147 additional CRC patients that were excluded from the survival analysis and found no differences in their PBL IGF2 methylation levels compared to those of the 281 CRC patients included in the survival analysis (**Supplementary File 5)**.

## 4. DISCUSSION

In this study, we first assessed the impact of the PBL IGF2 methylation status on the risk and prognosis of CRC. We found that individuals with IGF2 hypermethylation in their PBLs were at significantly higher risk of developing CRC than those with IGF2 hypomethylation. However, our findings showed better survival rates in CRC patients with PBL IGF2 hypermethylation compared to those in CRC patients with IGF2 hypomethylation.

In our initial case-control study, it is not possible to determine the aetiologically relevant time window of IGF2 methylation relative to CRC development. Fortunately, the prospective nature of the EPIC-Italy cohort is invaluable for confirming the temporal sequence of DNA methylation and CRC onset and therefore helps to distinguish causal from consequential epigenetic changes.[28 29]. In this nested case-control study, blood samples from CRC patients were collected 0.02-14.40 years (average 6.16 years) before diagnosis. Using this dataset, we found a positive but non-significant association between PBL IGF2 hypermethylation and an increased CRC risk. To account for reverse causality, we repeated the analyses excluding subjects who developed CRC within two years after their blood sampling and found that this positive association remained significant (PS-adjusted OR, 2.08, 95% CI: 1.18-3.69; *P*=0.012). Furthermore, our main finding was verified using the GSE89093 dataset. Finally, we pooled the results from these three datasets and found a 2.19-fold higher risk of developing CRC in the IGF2 hypermethylation group compared to the hypomethylation group.

The exact mechanism that links alterations in the IGF2 methylation status in PBLs and the susceptibility for CRC remains unclear. The PBL-derived DNA methylation profiles represent the overall methylation status of an individual. Alterations in PBL-derived DNA may be constitutively present before cancer occurs or represent an early response of the haematology system to the presence of tumour cells after cancer develops. Imprinting of IGF2 is primarily maintained by DNA methylation.[30 31] It has been reported that imprinting and expression are controlled by CpG-rich regions upstream of the IGF2 promotors. Normally, IGF2 is expressed from the paternal allele only, while the maternal allele is methylated and imprinted. Several previous studies have reported that the loss of imprinting (LOI) of IGF2 in peripheral blood (leukocytes or lymphocytes) may be a potential biomarker for the risk of developing CRC, [12 32] and IGF2 methylation alterations were suggested as a surrogate marker for LOI of IGF2.[32 33] If IGF2 aberrant methylation is merely a surrogate marker for LOI of IGF2, then IGF2 hypermethylation would be expected to be beneficial for maintaining the imprinting status of IGF2 and thus should decrease the risk of developing CRC. On the contrary, our results showed that IGF2 hypermethylation increases the risk of developing CRC, indicating that IGF2 participates in CRC tumorigenesis through two different modes of epigenetic alteration, aberrant hypermethylation and LOI, which is supported by previous studies.[34 35] Thus, our findings demonstrate that aberrant IGF2 hypermethylation can be assayed with a blood-based test and that the PBL IGF2 methylation status is likely to be a valuable predictive biomarker for CRC risk, independent of LOI of IGF2.

Further analysis of the GEO and TCGA datasets provided additional evidence supporting the association between IGF2 hypermethylation and the risk of developing CRC. For example, using these datasets we found significant associations between IGF2 hypermethylation in tissues and an increased risk of CRC or adenomas (**Figures 8A-D**). Interestingly, colorectal adenomas, a precancerous condition, showed similar levels of IGF2 methylation compared to tumour tissues (**Figure 8E**). Most colorectal cancers initially develop as benign precursor lesions (adenomas) that can take as long as 10 to 15 years to develop into carcinomas, which underpins early detection and removal of adenomas as an important strategy for preventing CRC.[6 36] The findings from the GEO and TCGA datasets indicate that IGF2 methylation in tissues can discern colorectal adenomas or CRCs from normal intestinal mucosa, which also suggests that IGF2 methylation may prove valuable during CRC screening for early cancer detection. We were unable to evaluate the association between IGF2 methylation in PBLs and the risk of developing adenomas. This issue should be explored in future studies.

**Figure 8.**
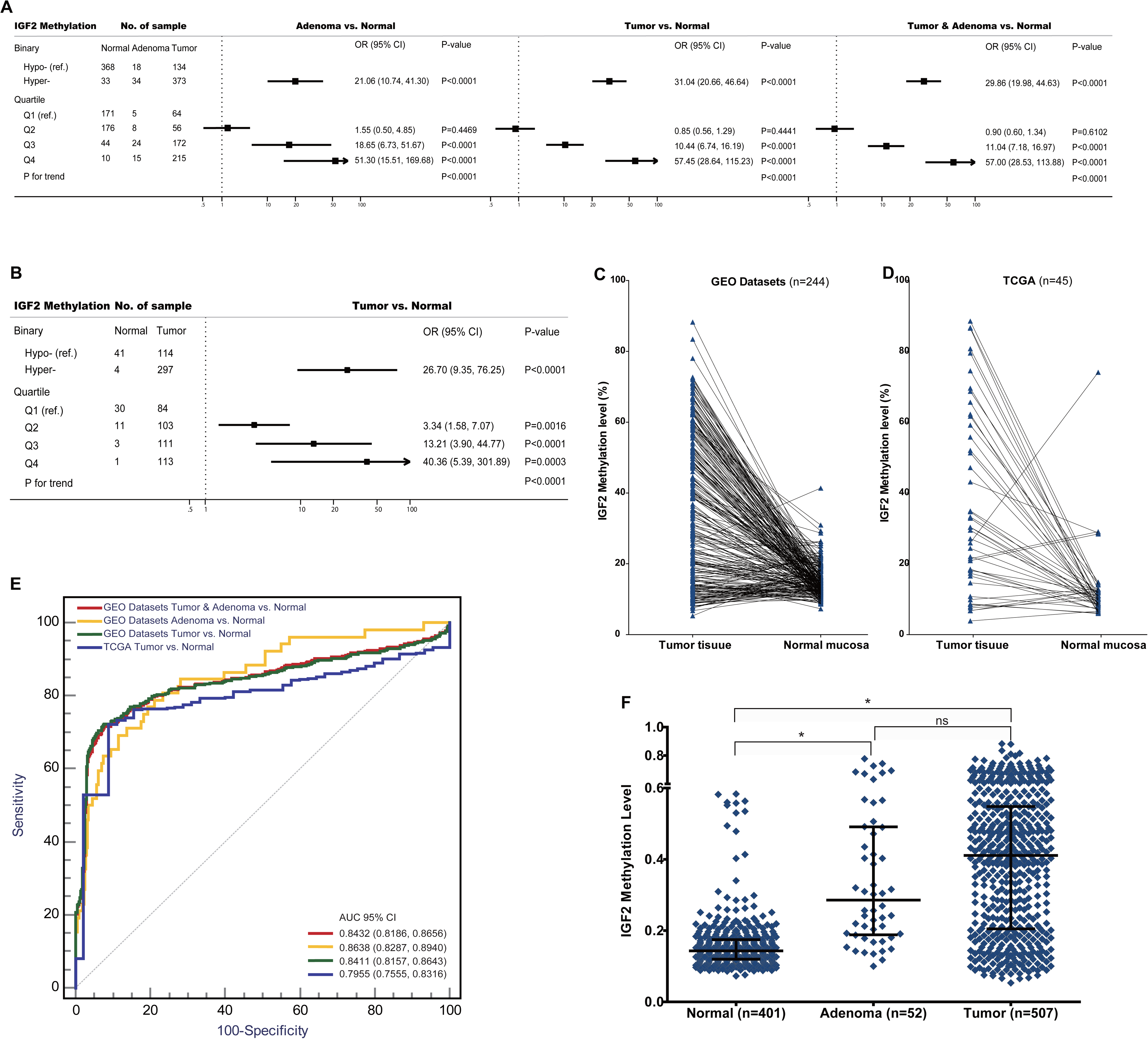
The associations between IGF2 methylation in tissue samples and CRC risk in the datasets from GEO and TCGA. (**A**) Adenoma or tumour tissues versus normal colorectal mucosa in the GEO datasets. (**B**) Tumour tissues versus normal colorectal mucosa in the TCGA datasets. (**C**) Tumour tissues versus matched adjacent normal tissues in the datasets from (**C**) GEO and (**D**) TCGA. (**E**) The discrimination performance of IGF2 methylation in tissue samples to identify CRC tumour or adenoma tissue and normal colorectal mucosa in the datasets from GEO and TCGA. (**F**) The IGF2 methylation levels in normal colorectal mucosa, adenoma and tumour tissue samples in the GEO datasets. *indicates *P*<0.0001, and ns indicates not statistical significance. AUC, the area under receiver operating characteristic curves; CI, confidence interval; OR, odds ratio; Hyper-, IGF2 hypermethylation; Hypo-, IGF2 hypomethylation;

In 2008, Ito and colleagues used peripheral blood samples to assess six CpG sites located in the IGF2 gene and reported that IGF2 methylation is not statistically associated with CRC risk.[34] However, the region tested in Ito’s study is different from the region examined in our study. As shown in **Figure 2**, the differentially methylated region assessed in our study is located near the TSS. To further validate our findings, we used the EPIC-Italy dataset to evaluate the methylation status of a relatively large region of −1500 bp upstream to +1500 bp downstream from the TSS. This sensitivity analysis again showed a statistically significant association between IGF2 hypermethylation and CRC risk (OR, 1.75, 95% CI: 1.15-2.66; *P*=0.009).

For the first time, we assessed the prognostic value of PBL IGF2 methylation status in CRC patients and found that patients with IGF2 hypermethylation in PBLs had significantly improved survival compared to patients with IGF2 hypomethylation. These findings were especially obvious for stage I-III CRC patients. However, this association did not reach statistical significance in the external EPIC-Italy CRC cohort. Given these inconsistent results, further cohorts with large sample size are needed to validate this novel finding.

For sensitivity analysis, we also evaluated the association between PBL IGF2 methylation and CRC prognosis during different follow-up periods. We observed a significantly strong association for 5-year relative survival, while the associations for both 1-year and 3-year relative survival were not statistically significant. Based on these results, it is hypothesised that the beneficial effects of IGF2 hypermethylation on CRC survival might begin the fifth year following CRC diagnosis and might persist for several years. In fact, the potentially beneficial effects may occur as early as the first year after CRC diagnosis, even though this effect did not reach statistical significance. The limited sample size of the cohort study may hinder the interpretation of these results; therefore, larger cohort studies are required to further evaluate this issue.

We also evaluated the impact of IGF2 methylation in tumour tissues on CRC patient survival and found no statistically significant association in both our initial cohort and the TCGA datasets. This is in consistent with a recent research.[7] However, two previous studies from Japan have assessed the IGF2 methylation status in tumour tissues of CRC patients and the results were inconsistent.[13 37] The region tested in those two studies is different from that examined in our study.

The lack of consistency between the results from the PBL samples and tumour tissues may reflect the fact that IGF2 methylation is merely a predictive marker rather than a prognostic marker. In addition, the detection of tissue-based markers depends on material from a biopsy or tumour tissue from resection. Because of intratumour heterogeneity, the detection of a biomarker from a single biopsy or one section of a tumour tissue sample might not necessarily represent the IGF2 methylation status of a given patient. Repeated biopsies and tests of multiple samples, however, are not feasible in routine clinical practice. Fortunately, blood-derived biomarkers have the potential to overcome these problems. In this respect, repeat blood sampling and detection of PBL IGF2 methylation is more acceptable and feasible than repeat biopsies in the clinic. Given that CRC has a wide range of long-term outcomes, PBL IGF2 methylation, as a DNA-based non-invasive blood test, could prove beneficial during follow-up and help identify patients at high risk of disease recurrence and progression.

An important aspect and potential concern of using PBL DNA methylation as a biomarker is whether leukocyte subpopulations affect the methylation signature of an individual. To address this concern, we collected patient clinical records including leukocyte counts and included these data as covariates in the PS model. Theoretically, the PS adjustment would control for the potential impact of different leukocyte counts and different subpopulations on our results. We compared the results before and after including the leukocyte count data in the PS model and found similar results (**Supplementary File 6**), suggesting that the effect of leukocyte counts and subpopulations on our results is negligible. Additional evidence supporting these findings can be found in several recently published studies which also showed that differences in leukocyte subpopulations were unlikely to interfere with the results of PBL-derived DNA methylation.[38 39]

Interestingly, our data indicates that PBL IGF2 hypermethylation correlates with serum CEA levels before surgery. Therefore, we further assessed whether the association between PBL IGF2 methylation and CRC prognosis is impacted by CEA levels. Using the additive model, we found a positive interaction between PBL IGF2 methylation and CEA on CRC prognosis (**Figure 9**). Given the limited sample size of the subgroups, this aspect should be further validated in future studies. Of note, after adjustment for CEA and the interaction between IGF2 methylation and CEA, the effect of IGF2 hypermethylation itself on CRC prognosis remained statistically significant (HR, 0.44, 95% CI: 0.24-0.79; *P*=0.006), suggesting a robust and independent role for PBL IGF2 hypermethylation in predicting the prognosis of CRC.

**Figure 9.**
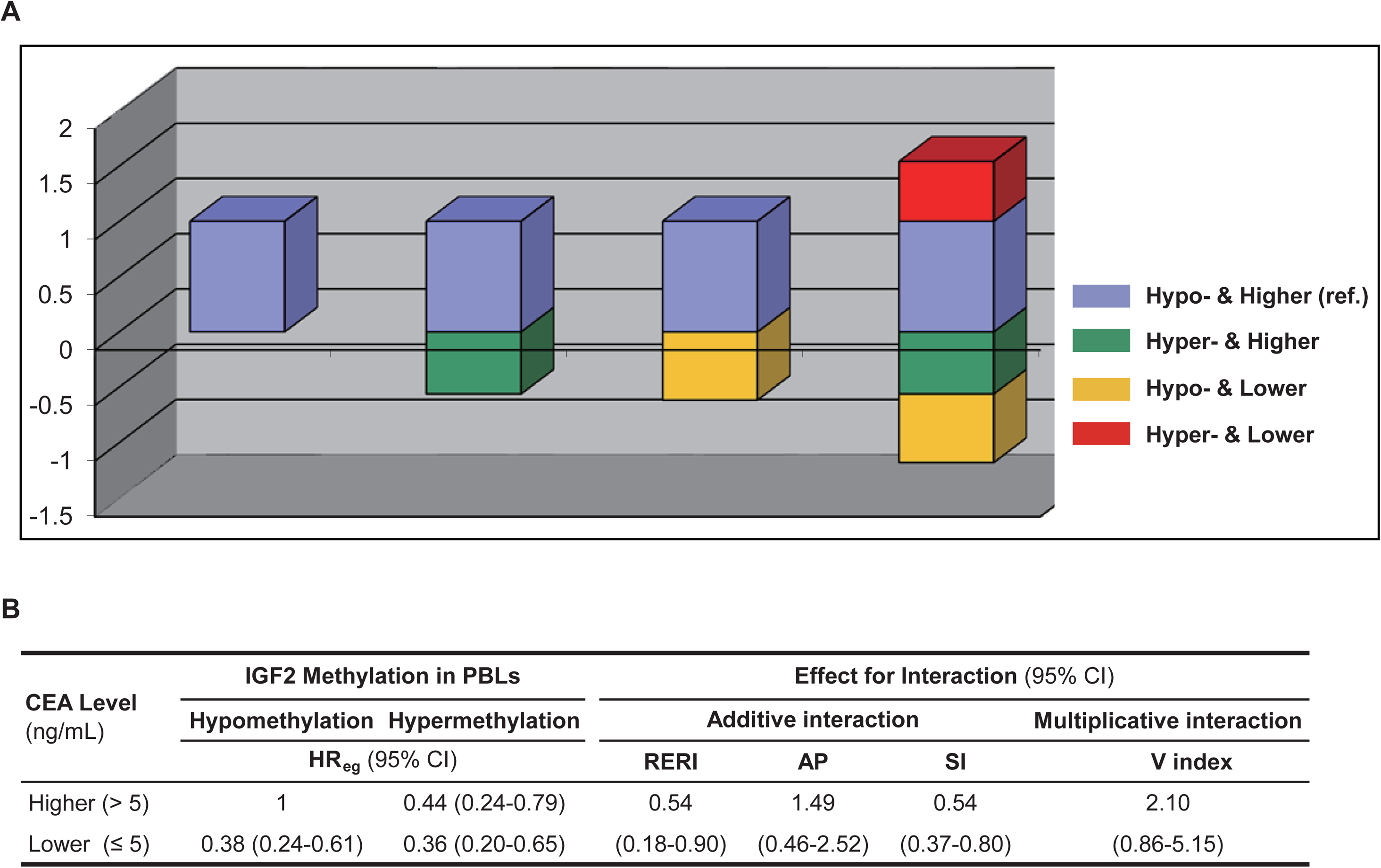
The interaction between PBL IGF2 methylation and serum CEA levels on CRC patient prognosis using the additive and multiplicative models. (**A**) The interaction analysed with the additive model. (**B**) The effect estimates for interaction in respected to both the additive and multiplicative models. AP, attributable proportion due to interaction; CI, confidence interval; HR, hazard ratio; Hyper-, IGF2 hypermethylation; Hypo-, IGF2 hypomethylation; PBL, peripheral blood leukocyte; RERI, relative excess risk due to interaction; SI, synergy index; V index, the multiplicative interaction index.

This study has several strengths. The findings from our initial studies were validated with several external datasets. In our initial CRC patient cohort, the covariates were collected prospectively and were blinded to patient outcome. We used PS techniques to control for multiple potential confounding factors. Furthermore, we performed extensive sensitivity analyses to assess the robustness of our findings. The confounding RR and the E-value sensitivity analyses demonstrated that our results are unlikely to be substantially impacted by both the adjusted confounders included in the PS models or a potential residual confounder.

In our present cohort, adjuvant chemotherapy was not offered routinely to high risk individuals. Thus, we did not analyse the clinical significance of IGF2 methylation as a predictive biomarker for sensitivity to adjuvant chemotherapy. Further studies are needed to explore and clarify this issue. Another potential limitation is the limited sample size used in the stratified analyses. Therefore, the results from the stratified analyses should be interpreted with caution.

## 5. CONCLUSIONS

In summary, IGF2 methylation in PBLs is significantly associated with the risk and prognosis of CRC, suggesting an important role for IGF2 methylation as a blood-based predictive biomarker to identify of individuals at high risk of developing CRC; meanwhile, PBL IGF2 methylation might also serve as a predictive biomarker for CRC prognosis.

## Data Availability

Authors are willing to share any data that are used in this work. The data that support the findings of the initial study are included in the manuscript and supporting file. The raw datasets used in the validation stage are publicly available on GEO (http://www.ncbi.nlm.nih.gov/geo/) and TCGA (CORD and READ) (https://cancergenome.nih.gov/).

## Abbreviations

body mass index (BMI), carcinoembryonic antigen (CEA), confidence intervals (CIs), colorectal cancer (CRC), CRC-specific survival (CSS), disease free survival (DFS), Gene Expression Omnibus (GEO), Harbin Medical University (HMU), hazard ratios (HRs), insulin-like growth factor 2 (IGF2), loss of imprinting (LOI), methylation-sensitive high-resolution melting (MS-HRM), odd ratios (ORs), overall survival (OS), peripheral blood leukocytes (PBLs), propensity score (PS), The Cancer Genome Atlas (TCGA).

## Acknowledgments

The authors thank the participants and staff of both the initial study and the validation studies from the GEO and TCGA for providing data and their valuable contributions. The authors thank the GEO and TCGA databases for the access to the validation datasets. The authors thank Dr. Carlotta Sacerdote (Città della Salute e della Scienza University) and Dr. Giovanni Fiorito (University of Sassari) for providing critical help in validation stage. The authors thank Professor Wei Chen (Tulane University) for critically reading this manuscript.

## Supplementary Files

Supplementary File 1A. Main characteristics of participants of the initial and validation studies in the CRC risk analysis.

Supplementary File 1B. Main characteristics of patients of the initial and validation cohorts in the CRC prognosis analysis.

Supplementary File 2A. Comparisons of participant characteristics and covariates between CRC cases and controls before and after propensity score adjustment in the initial case-control study in CRC risk analysis.

Supplementary File 2B. Comparisons of baseline characteristics of CRC patients before and after propensity score adjustment in the initial cohort in CRC prognosis analysis.

Supplementary File 3A. Sensitivity analyses using additional propensity score based methods for CRC risk analysis in the initial case-control study.

Supplementary File 3B. Sensitivity analyses using additional propensity score based methods for CRC survival analysis in the initial CRC cohort.

Supplementary File 4. E-values for PS-adjusted effect estimates.

Supplementary File 5. Associations between PBL IGF2 methylation status and UICC stage, serum CEA level, WBC counts, and whether or not included in the survival analysis among CRC patients.

Supplementary File 6. Associations of PBL IGF2 methylation and CRC patient prognosis with or without WBC counts included in PS models.

